# An optimized stepwise algorithm combining rapid antigen and RT-qPCR for screening of COVID-19 patients

**DOI:** 10.1101/2021.01.13.21249254

**Authors:** Philippe Halfon, Guillaume Penaranda, Hacène Khiri, Vincent Garcia, Hortense Drouet, Patrick Philibert, Christina Psomas, Marion Delord, Julie Allemand-Sourrieu, Frédérique Retornaz, Caroline Charpin, Thomas Gonzales, Hervé Pegliasco, Jérôme Allardet-Servent

## Abstract

**Background:** Diagnosing SARS CoV-2 infection with certainty is essential for appropriate case management. We investigated the combination of rapid antigen detection (RAD) and RT-qPCR assays in a stepwise procedure to optimize the detection of COVID-19.

**Methods:** From August 2020 to November 2020, 43,399 patients were screened in our laboratory for COVID-19 diagnostic by RT-qPCR using nasopharyngeal swab. Overall, 4,691 of the 43,399 were found to be positive, and 200 were retrieved for RAD testing allowing comparison of diagnostic accuracy between RAD and RT-qPCR. Cycle threshold (Ct) and time from symptoms onset (TSO) were included as covariates.

**Results:** The overall sensitivity, specificity, PPV, NPV, LR-, and LR+ of RAD compared with RT-qPCR were 72% (95%CI 62%–81%), 99% (95% CI95%–100%), 99% (95%CI 93%–100%), and 78% (95%CI 70%–85%), 0.28 (95%CI 0.21-0.39), and 72 (95%CI 10-208) respectively. Sensitivity was higher for patients with Ct ≤ 25 regardless of TSO: TSO ≤ 4 days 92% (95%CI 75%–99%), TSO > 4 days 100% (95%CI 54%–100%), and asymptomatic 100% (95%CI 78-100%). Overall, combining RAD and RT-qPCR would allow reducing from only 4% the number of RT-qPCR needed.

**Conclusion:** This study highlights the risk of misdiagnosing COVID-19 in 28% of patients if RAD is used alone. Thus, negative results from RAD needs to be confirmed by RT-qPCR prior to making treatment decisions. A stepwise analysis that combines RAD and RT-qPCR would be an efficient screening procedure for COVID-19 detection and may facilitate the control of the outbreak.

## Introduction

Since the beginning of the COVID-19 outbreak, one of the most important challenges for the scientific community is to quickly establish the diagnosis of SARS-CoV-2 infection. To establish an effective COVID-19 filter that will stop this pandemic, tests that can enable both mass screening and reliable detection of infected people are needed^1^. Consequently, the use of surveillance testing to identify infectious individuals represents one possible method for breaking enough transmission chains in order to suppress the ongoing pandemic and reopen societies with or without a vaccine. The reliance on testing underscores the importance of analytical sensitivity of virus assays with gold-standard being the real-time quantitative polymerase chain reaction (RT-qPCR) from nasopharynx swab^2^.

These assays have analytical limits of detection that are usually within 10^3^ viral RNA copies per ml (cp/ml)^3,4^. However, RT-qPCR remains expensive and often has sample-to-result times of 24–48 h as a laboratory-based assay. New developments in SARS-CoV-2 diagnostics have the potential to reduce cost significantly, thus allowing for expanded testing or greater frequency of testing and reducing turnaround time to minutes^5–13^. Several diagnostic strategies are available for identifying or ruling out current infection, identifying people in need of care escalation, or testing for past infection and immune response. Point-of-care (POC) antigen and molecular tests for the detection of current SARS-CoV-2 infections could allow for the earlier detection and isolation of confirmed cases than laboratory-based diagnostic methods, with the aim of reducing household and community transmission. These tests exist today in the form of rapid molecular assays or rapid antigen detection (RAD) tests. The latter can quickly detect fragments of proteins found on or within the virus by testing samples collected from the nasal cavity via swabs. Additionally, RAD tests, which are widely used to detect viral infections other than COVID-19, are not only rapid (15–30 minutes) but also simple to use, require short training, and can be performed outside the laboratory for mass screening when RT-qPCR assays are not or insufficiently available. RAD tests are cheap (<5 USD), can be produced in tens of millions or more per week; these advantages can theoretically lead towards effective COVID-19 filter regimens. However, these assays do not meet the gold standard for analytical sensitivity, thus hindering their widespread use ^14^. Among factors that potentially alter the sensibility of RAD tests are the viral load, which has been found to be highly variable in COVID-19 patients and depends on factors such as time from symptoms onset (TSO), sample collection (i.e., type and quality), disease severity, and patient age^14–16^. Zou et al.^14^ reported cycle threshold (Ct) values in the range of 19–40 in the upper respiratory specimens of infected patients. Antigen tests are less sensitive than RT-qPCR and could be less reliable in the clinical diagnosis of COVID-19 patients with low viral load. Caraguel et al.^17^ reported that epidemiologic strategies use criteria based on either the probability or the cost of a false test result associated with a specified cutoff. Recent studies on four different commercial RAD tests demonstrated a wide range of sensitivities from 16.7% to 85% (with 100% specificity) in COVID-19 clinical samples^12,18,19^. World Health Organization recommends RAD tests that meet the minimum performance requirements of ≥80% sensitivity and ≥97% specificity, while the European Centre for Disease Prevention and Control suggests to use tests with a performance closer to those of RT-PCR, i.e. ≥90% sensitivity and ≥97% specificity^20,21^. The objective of this study was to assess the combination of RAD and RT-qPCR assays in a stepwise procedure to optimize the detection of COVID-19 in a large cohort of patients.

## Patients and Methods

From August 2020 to the beginning of November 2020 in Marseille (i.e., during the second wave of the COVID-19 outbreak in France), COVID-19 diagnostic tests were assessed by RT-qPCR in 43,399 patients using nasopharyngeal samples. The symptoms and presumed time from symptoms onset (TSO) were recorded. Overall, 4,691 (11%) of the 43,399 were found to be positive for COVID-19 and 38,708 negatives (89%). Among COVID-19 negative patients, TSO was ≤4 days in 4,836 (12%), >4 days in 2,134 (6%), 26,800 were asymptomatic (69%), and 4,938 (13%) did not know when symptoms appeared. Among the 4,691 COVID-19 positive patients, TSO was ≤4 days in 1,629 (35%), >4 days in 756 (16%), 1,684 (36%) were asymptomatic, and 621 (13%) did not know when symptoms appeared (and one patient did not answer the question). The Ct values of RT-qPCR positive samples according to TSO are shown in study flowchart (Figure 1) and in Figure 2. Considering the distribution of both TSO and Cts, 200 samples were also tested by RAD, among whom 100 samples (50%) were positive for COVID- 19 and 100 samples (50%) were negative for COVID-19 (Figure 1).

**Figure 1:**
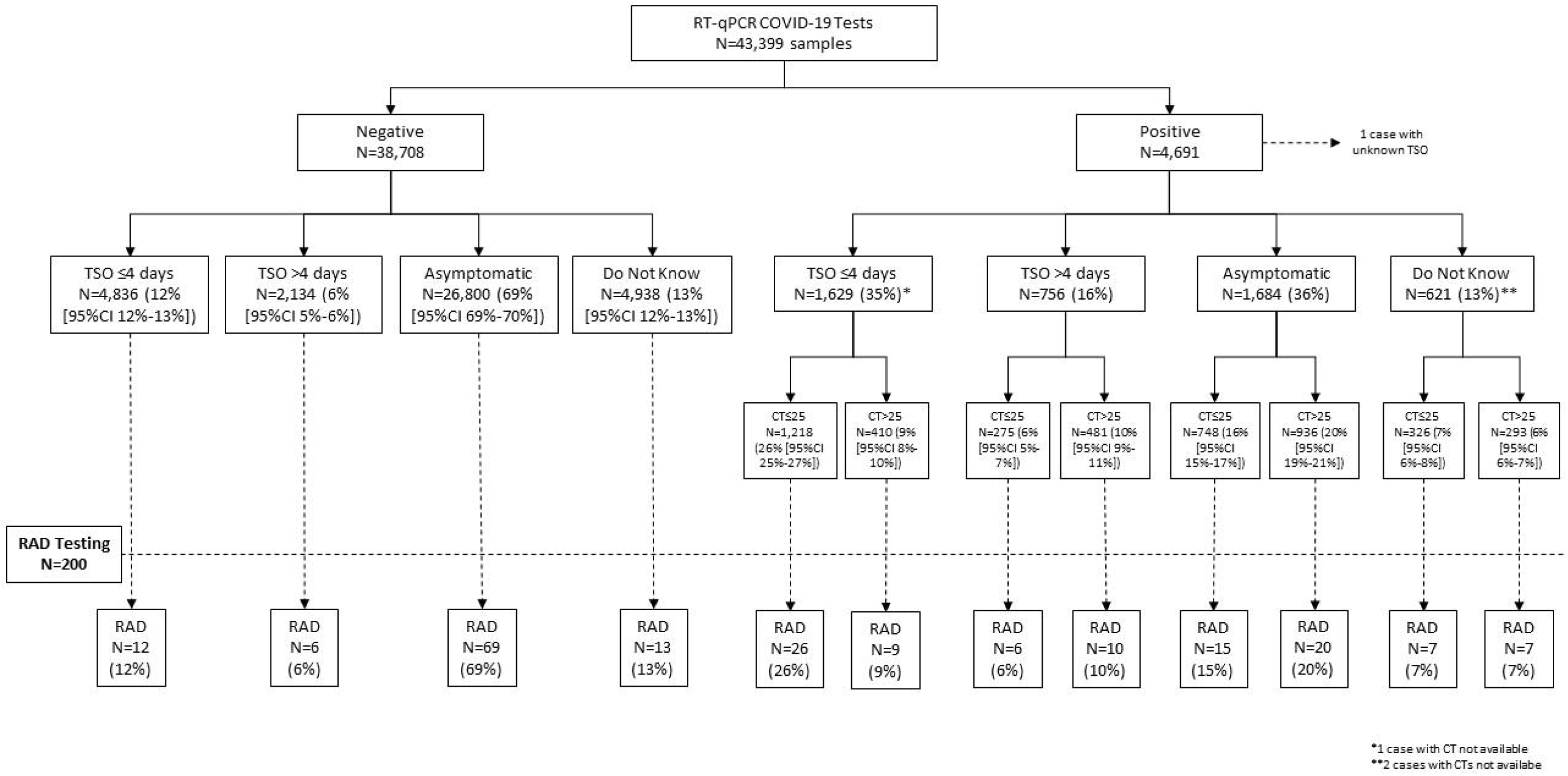
Study Flowchart. TSO, time from symptoms onset; RAD, rapid antigen detection

**Figure 2:**
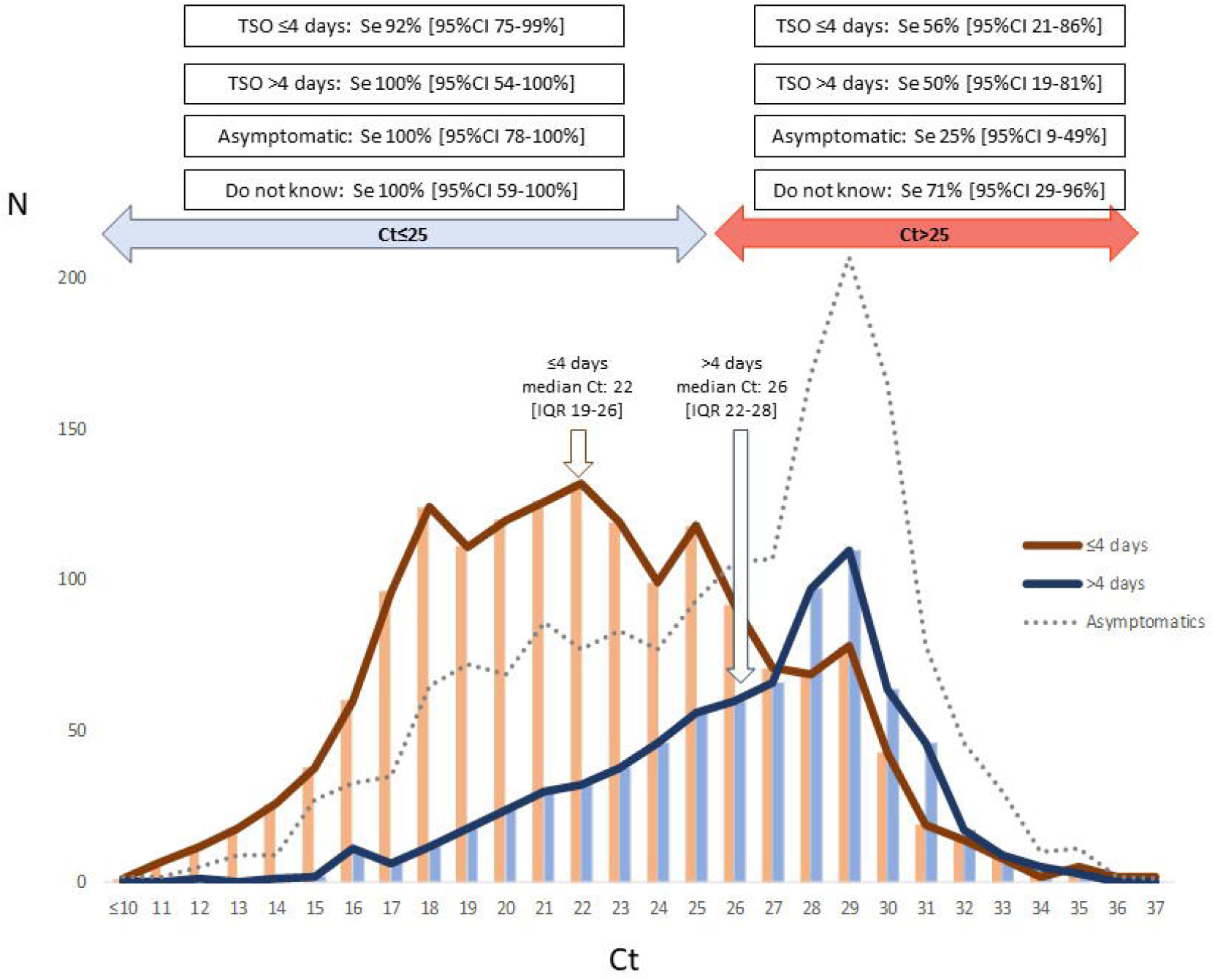
Ct values according to time from symptoms onset, and sensitivities of rapid antigen detection assay according to Ct and time from symptoms onset. TSO, time from symptoms onset

RT-qPCR testing was performed under routine conditions by using the UltraGene Combo2Screen SARS-CoV-2 Assay (ref 139b) (ABL SA Group, Luxembourg) and Chemagic™ viral DNA/RNA 300 kit H96 (ref. CMG-1033-S) on a Chemagic™ 360-D instrument (PerkinElmer, Inc., Austin, TX) according to the manufacturer’s instructions^22^. RAD testing was performed using a Panbio™ COVID-19 Ag Rapid Test Device according to the manufacturer’s instructions^23^.

According to French regulations, the study was submitted to French ethics committee (CPP Sud-Méditerranée II) and registered as a reference methodology (MR-004) on the Health Data Hub French registering website platform (registration number: F20201028125903, https://www.health-data-hub.fr). Patients were informed that their biological results could be used for research purposes and that they were free to refuse participation in the study.

Sensitivity, specificity, positive predictive value (PPV), negative predictive value (NPV), negative likelihood ratio (LR-), and positive likelihood ratio (LR+) were reported using frequencies and percentages with exact 95% CI. Fisher’s exact test was used to compare the sensitivity of RAD controlling for Ct (≤ 25 and > 25). Comparisons of stratified groups were performed using Cochran-Mantel-Haenszel Chi-Squared test. Statistical significance was assumed at p < 0.05. Calculations were performed using SAS V9.4 software (SAS Institute Inc., Cary, NC, USA).

## Results

### RAD sensitivity

Among the 200 patients tested by RAD, there were 104 females (52%) and 96 males (48%); mean age was 48 years old (standard deviation, 21). The overall sensitivity, specificity, PPV, NPV, LR-, and LR+ of RAD compared with RT-qPCR were 72% (95% CI 62%–81%), 99% (95% CI 95%–100%), 99% (95% CI 93%–100%), 78% (95% CI 70%–85%), 0.28 (95% CI 0.21-0.39), and 72 (95% CI 10-208) respectively. The sensitivity of RAD according to Ct values was significantly higher in Ct ≤ 25 than in Ct > 25: 96% (95% CI 87%–100%) vs. 44% (95% CI 29%–59%), respectively (p < .0001). Sensitivity was even lower in CT > 30: 20% (95% CI 3%–56%). The sensitivity of RAD according to TSO was 83% if TSO ≤ 4 days from RT-qPCR (95% CI 66%–93%), 69% if TSO > 4 days from RT-qPCR (95% CI 41%–89%), and 57% in asymptomatic patients (95% CI 39%–74%) (p = 0.0661).

The interactions between Ct values and TSO showed a predictably higher sensitivity for Ct ≤ 25 regardless of TSO: TSO ≤ 4 days 92% (95% CI 75%–99%), TSO > 4 days 100% (95% CI 54%–100%), and asymptomatic 100% (95% CI 78-100%). For patients with Ct > 25, sensitivity was higher when TSO ≤ 4 days than when TSO > 4 days or even in asymptomatic patients but was still not significant: 56% (95% CI 21%–86%), 50% (95% CI 19%–81%), and 25% (95% CI 9%-49%)respectively (p = 0.2099) (Figure 2).

### Combination of RAD and RT-qPCR

Figure 3 shows a stepwise algorithm that combines RAD and RT-qPCR for the screening of COVID-19 patients. As sensitivity of RAD is over 80% only in patients with TSO ≤4 days (i.e., 83%), RAD might be useful only in such patients. If RAD was used in first intention among the 6,465 patients with TSO ≤4 days, a COVID-19 positive results would be expected in 1,739 patients (4% of overall 43,399 COVID-19 diagnostic tests assessed by RT-qPCR) : 1,629 patients with TSO ≤4 days are COVID-19 positive by RT-qPCR and sensitivity of RAD is 83% among these patients, and 4,836 patients with TSO ≤4 days are COVID-19 negative by RT-qPCR and specificity of RAD is 92% among these patients (i.e., estimated 8% false positive cases). Thus, a confirmatory RT-qPCR test would be required in the 4,726 patients with a COVID-19 negative RAD result.

**Figure 3:**
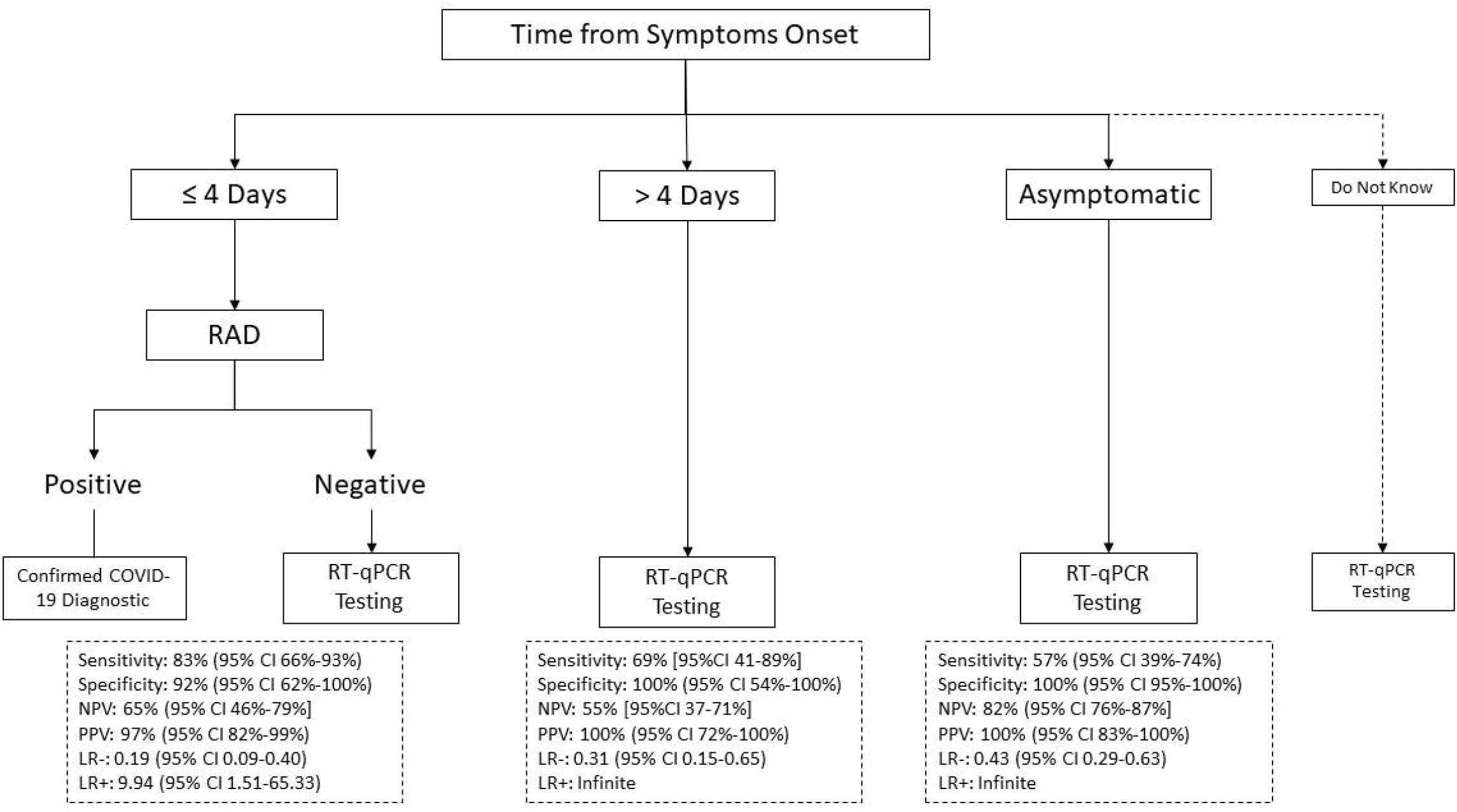
Stepwise algorithm combining rapid antigen detection and RT-qPCR. PPV, positive predictive value; NPV, negative predictive value; LR-, negative likelihood ratio; LR+, positive likelihood ratio

Overall, RAD would be used in 6,465 patients, and RT-qPCR would be used in 41,660 (28,484 asymptomatic, 2,890 TSO >4 days, 5,560 patients who do not know TSO, and the 4,726 confirmatory RT-qPCR in COVID-19 negative RAD results). Using RAD in 6,465 patients among all 43,399 patients attending the laboratory for COVID-19 diagnostic would allow reducing from only 4% the number of RT-qPCR needed.

## Discussion

In this study, we propose an optimized stepwise analysis that combines RAD and RT-qPCR for the screening of COVID-19 patients. This study, performed in a large cohort of 43,399 patients in real life conditions, highlights the risk of misdiagnosing COVID-19 in 28% of 4,691 patients (i.e., n=1,314) if RAD was used alone for diagnosis. A large implementation of RAD in the current surveillance approach can lead to the identification of infected people when the test is positive. However, this approach is slow, thus leading to the potential spread of infection when the test is negative. This limits the impact of isolation and contact tracing. If RAD has a sensitivity of at least 80%, it might be used only in in patients with TSO ≤ 4 days and need to be confirmed by an RT-qPCR assay when negative.

One of the main issues that the scientific community have to deal with when using RAD as a mass screening process is its low sensitivity (30%–75%) depending on the assays and the type of population analyzed; this issue has been described by several teams^2^. In a recent Cochrane study, antigen test sensitivity varied considerably across studies (from 0%–94%): the average sensitivity and average specificity were 56.2% (95% CI 29.5%–79.8%) and 99.5% (95% CI 98.1%–99.9%), respectively (based on 8 evaluations in 5 studies on 943 samples)^24^. Data for individual antigen tests were limited, with no more than two studies for any test^24^. Rapid molecular assay sensitivity showed less variation than antigen tests (from 68% to 100%), with an average sensitivity of 95.2% (95% CI 86.7%–98.3%) and average specificity of 98.9% (95% CI 97.3%–99.5%) (based on 13 evaluations in 11 studies of 2,255 samples).

A high sensitivity for SARS-CoV-2 detection in nasopharynx swab samples was observed only for samples with Ct < 25 (corresponding to viral loads higher than 10^6^ copies/mL, which has been proposed as the threshold of transmissibility)^17^. However, the reported sensitivities were lower and more variable (72.2%, 95% CI 49.1%–87.5%) in samples with low viral load (Ct > 25.1). Like molecular tests, antigen tests are typically highly specific for the SARS-CoV-2 virus. However, all diagnostic tests may be subject to false-positive results, particularly in low prevalence settings with false-negative results. Healthcare providers should always carefully consider diagnostic test results in the context of all available clinical, diagnostic, and epidemiological information. Health care providers and clinical laboratory staff can help ensure the accurate reporting of test results by following the authorized testing instructions and key steps in the testing process recommended by the Centers for Disease Control and Prevention, including routine follow-up testing (reflex testing) with a molecular assay when appropriate, and by considering the expected occurrence of false-positive results when interpreting the test results in their patient populations. More ideal POC sample types, such saliva, are less invasive, and their adoption is expected to accelerate the use of antigen tests as a much-needed screening tool. A recent evaluation reported a low sensitivity of 11.7% for self-collected COVID-19 positive saliva samples when using antigen tests^19^. Given the increasing global need for COVID-19 tests, rapid and inexpensive assays are required to supplement current nucleic acid amplification–based assays, and the wide variation in the sensitivities of RAD needs to be evaluated and understood.

Regardless of whether 72% of the overall sensitivity of the RAD (PANBIO COVID-19) is in agreement with those found by Fenollar et al.^25^, the scientific community has to pay attention to the panel of nucleic acid amplification testing positive controls used for testing the sensitivity of RAD. We showed that sensitivity varies from 25% to 100% according to TSO and Cts (≤25, >25).

Based on our experience and others, we would consider using RAD only for patients with TSO≤ 4 days (sensitivity: 83% [95% CI 66%–93%]) if the test is positive, when RT-qPCR is unreliable, and when RT-qPCR cannot be reported in a short time (<24 h). Otherwise, any negative RAD should be confirmed by an RT-qPCR assay. This timing of TSO ≤ 4 days is in agreement with the fact that the RT-qPCR false-negative rate is the lowest three days after the onset of symptoms or approximately eight days after exposure^26^. Clinicians should consider waiting one to three days after the onset of symptoms to minimize the probability of a false-negative result. Notably, we found that age (>65 or <65 years) had no effect on the decision to use RAD first or RT-qPCR first (results not shown). If the availability of POC or self-administered surveillance tests leads to a faster turnaround time or more frequent testing, our results suggest that they would have a high epidemiological value. We showed that using RAD may allow reducing from only 4% the number of RT-qPCR performed. One of the major advantages of RAD in the effectiveness of surveillance of the outbreak beside shorter turnaround time and lowest cost is its speed of reporting more than its sensitivity^27^

Our study has some limitations. First, we do not assess RAD in the whole population of 43,399 patients but RAD was performed on a sample representative for TSO and Cts distribution among all RT-qPCR samples. Another limit is that calculations were performed on samples among whom 11% were positive for COVID-19; thus, the usefulness of RAD must be re- assessed according to the prevalence of COVID-19 by RT-qPCR.

In summary, surveillance should prioritize sensibility, accessibility, frequency, and sample-to-answer time. However, based on the current understanding of sensitivity challenges, our study may alert the scientific community to the fact that extensive use and misinterpretation of RAD can lead to the misdiagnoses of COVID-19 patients due to its low predictive negative value. Negative results from an antigen test should be considered in context of the clinical observations, patient history, and epidemiological information and may need to be confirmed with a molecular test prior to making treatment decisions. A stepwise analysis that combines RAD and RT-qPCR would be an efficient screening procedure for COVID-19 detection and may facilitate the control of the outbreak.

## Data Availability

Data will be available immediately after publication with no end date to researchers who provide a methodologically sound proposal. Requests should be addressed by email to.

